# Feasibility of Community Health Worker based cardiovascular risk reduction strategies in urban slums of Bhopal: Rationale, design and baseline results of community based study

**DOI:** 10.1101/2020.09.18.20189639

**Authors:** Abhijit Pakhare, Ankur Joshi, Sagar Khadanga, Sanjeev Kumar, Shubham Atal, Vaibhav Ingle, Yogesh Sabde, Neelesh Shrivastava, Anuja Lahiri, Akash Ranjan, Rajnish Joshi

## Abstract

In urban India, about 35-40% of all adults have hypertension and about 10-15% have diabetes mellitus. National Program for Prevention and Control of Diabetes, Cancer and Stroke (NPCDCS) launched by Government of India has envisaged to screen all adults aged 30 years and above for presence of hypertension and diabetes mellitus in the community through population-based screening, initiate positively screened and diagnosed on drug therapy, and follow them up for treatment adherence. In this context, the current study aims to estimate burden of high cardiovascular disease (CVD) risk and to evaluate feasibility of community health workers-based strategies in reducing CVD among adults living in urban slums. We have identified and trained CHWs from within the urban slum communities in Bhopal, in chronic disease identification, skills in self-care and adherence promotion skills. At-risk individuals were linked to public health facilities as outlined under NPCDCS. Primary outcome is assessment of burden of high cardiovascular risk and its determinants. Secondary outcome is feasibility of community health worker-based adherence promotion. Between November 2017 and June 2018, CHWs in 14 urban slum clusters, screened a total of 6178 individuals out of which 4781 (77.43%) attended confirmation camp. Around 2393 (38.8%) were current tobacco users (smoking and/or smokeless), and 4697 (76.1%) has a sedentary lifestyle. Out of 758 (12.3%) known hypertensives, 354 (46.7%) had controlled hypertension whereas out of 333 (5.4%) known diabetes patients, 169 (57.5%) has controlled level of diabetes. In 813 (15%) out of 5416 and 151 (3.4%) out of 4486 adults, hypertension and diabetes was newly detected respectively. Results of this study have a potential to strengthen NPCDCS across all urban areas of the country. This manuscript describes detailed protocol of the study and presents baseline summary of CVD risk factor burden in urban slums of Bhopal.

## Introduction

Cardiovascular diseases (CVDs, which include coronary artery disease, cerebrovascular disease, and peripheral vascular disease) are the leading cause of morbidity and mortality among adults both in urban and rural India (1). In Madhya Pradesh (MP) the disease burden attributable to NCDs is 50.5% with CVDs as leading cause (2). Prevalence of CVD risk factors such as tobacco use, hypertension and diabetes is high in both urban and rural India with MP being no exception (3-7). Proportionate morbidity and mortality due to CVDs is greater in urban as compared to rural populations (8-10). Despite a lower risk-factor burden in low-income as compared to high-income countries, CVD mortality in low-income countries is high (11). This evidence implies that control of CVD risk factors remains poor in India (12,13).

Approximately 377 million individuals reside in urban India (2011 census; 430 million by 2021). About a quarter of all urban residents live in slums, a number which is projected to double in next 10 years. Urban slums are considered disadvantaged in many ways. While food, shelter and security are the primary concerns, health is not a priority and burden of disease is high. Limited data indicate that CVDs are a significant problem. In a recent study among 2,564 residents of an urban slum in North India,(14) among men and women 46.7% and 9.9% used tobacco, 16% and 21.9% were overweight, prevalence of hypertension (cut-off 140/90) was about 16%. As per a study conducted in 2014, the hypertension prevalence in slums of Delhi was as high as 23% (15). Another study conducted on urban slum dwellers of Kolkata depicted the prevalence of diabetes as 13%, high BMI as 56%, smoking as 21% and obesity as 56% (16). The overall cardiovascular risks are greater in low socioeconomic strata in urban India (17). Clearly there is an urgent need to intervene.

Primary prevention interventions with an eventual aim to reduce CVD events have focussed on improving awareness, screening, and initiating individuals on evidence based effective therapies. Despite a documented high prevalence of CVD risk factors across the globe, control of risk factors remains a challenge in developing as well as developed world (18,19). These are human resource intensive tasks, which need a health-systems approach. The healthcare system in many developing countries including India is fractured, and overall resources are limited (20,21). The urban public health system is much less developed as compared to the rural areas and provides only limited CVD care. Although urban households are often in proximity to good public and private health care facilities, residents have restricted access due to social and economic reasons. The National Urban Health Mission (NUHM) as presently planned is in its infancy. Thus, there is an urgent need to develop evidence of feasible, effective, and low-cost interventions to control risk factors for NCDs in urban areas. Such an intervention, if beneficial can be later integrated into the existing public health delivery system.

A non-physician community health worker (CHW) based household level intervention has a potential to enhance knowledge, improve attitudes, and reduce risk factor levels for NCDs. CHW based healthcare delivery models have been shown to be effective in maternal and neonatal care, malaria, tuberculosis, and HIV. Evidence of their effect is however lacking in CVD and DM. The latter are considered to be complex, and often need a multi-dimensional approach, which may be beyond the capabilities of CHWs. Hence there is equipoise in the effectiveness of CHW based healthcare delivery model for NCDs.

Overall aim of this study is to estimate burden of high cardiovascular disease (CVD) risk and to evaluate feasibility of community health workers-based strategies in reducing CVD among adults living in urban slums.

## Primary Objectives

1. To assess prevalence of common modifiable risk factors (tobacco consumption, high salt consumption, physical inactivity, harmful use of alcohol, hypertension, diabetes, obesity, etc) for CVDs in urban slums
2. To identify and train community health workers to screen individuals at a high risk for future cardiovascular events, and impart skills of self-care, risk reduction strategies and assessment of treatment adherence.
3. To increase awareness about common modifiable risk factors using inter-personal communication.

## Secondary Objectives

1. To improve linkages of communities to public health delivery services for CVDs as outlined in National Program for Prevention and Control of Cancer, Diabetes, Cardiovascular Diseases and Stroke (NPCDCS).
2. To evaluate feasibility of community health worker-based adherence promotion strategies.

## Methods

### Design and Ethics Statement

We have designed a community-based study determine burden of CVD risk factors in urban slums and feasibility of CHW based adherence promotion activities. Burden of CVD risk factors was determined in a cross-sectional assessment of urban slum population. Those found at high risk were longitudinally followed up over next 18 months to understand impact of adherence promotion activities. The study design was approved by the institutional human ethics committee (Ref: IHEC-LOP/2017/EF00045) and funded by Indian Council of Medical Research. All participants were recruited only after obtaining written informed consent.

### Setting

Bhopal is the second largest city in state of Madhya Pradesh with its population of approximately 2 million according to 2011 census. Bhopal city is divided into 14 Zones and 85 wards. Wards are administrative units of Bhopal Municipal Corporation. Bhopal city has over 350 designated urban slums. The study was conducted in 16 urban slum clusters of Bhopal. Cluster is a sub-unit of a designated urban slum, consisting of about 250 households, and about 1000 individuals. There is one frontline community health worker known as ASHA (Accredited Social Health Activist) designated as facilitator for all public health programme implementation for such population. ASHA is usually a high-school educated female, who receives incentive-based payments for health-care delivery tasks she is trained to perform. While their initial training was to deliver reproductive and child health, under NPCDCS they are being trained for NCD screening activities. For the purpose of health-care delivery, a group of clusters form catchment area for a urban primary health center (UPHC), which is a public sector facility, that provides primary health. The community from which the study participants were selected in current study normally seek public health care at either *Barkheda Pathani* or *Sai-baba Nagar* UPHCs. There were no out-of-pocket costs towards either consultation or available medications at these facilities.

### Participants

The target population is *individuals aged 30 years or more living in an urban slum in Bhopal*, India. The target population will be identified in a door-to-door survey conducted by ASHA. Individuals residing in the selected clusters for at least last 6 months were eligible for inclusion. Women who are pregnant at the time of screening were excluded. No other exclusions were applied. Amongst this population, we have identified high risk group for CVD events as individuals with hypertension (previously known hypertension or SBP > 140 or DBP >90 mm Hg on two or more occasions), diabetes (Previously known diabetes or Random blood sugar > 200mg/dL or a fasting blood sugar > 126mg/dL), a previous known cardiovascular or a cerebrovascular event. This high-risk group was followed up over next eighteen months to demonstrate feasibility of CHW based interventions.

### Study Procedures

#### ASHA recruitment and training

ASHAs from all the selected clusters were approached by study supervisors who are medical social workers trained by the study investigators. The ASHAs willing to participate in the study were provided with training in key measurement skills. The training program covered knowledge regarding basic Health issues with respect to CVD, its risk factors, and their prevention. The training focused on key skills that include interview and Communication techniques, measuring physical parameters like Height, Weight, waist measurement, blood pressure measurement using a digital sphygmomanometer, and blood sugar estimation using a glucometer.

#### ASHA engagement and home-based screening

All ASHAs were asked to sub-divide their clusters into five segments consisting of 50 households each. The ASHAs made a microplan for home-based screening guided by study supervisors. ASHA were provided a performance-based incentive contingent on confirmation of screening by the study supervisor. ASHA administered a screening questionnaire (to identify tobacco or alcohol consumption, previously known hypertension, diabetes or a manifest CVD such as ischemic heart disease or stroke), then performed anthropometry (to measure body mass index (BMI) and waist circumference (WC) (weighing scale -Seca-876, stadiometer-Seca-213 and measuring tapes Seca-201, Seca, Hamburg, Germany), measured blood-pressure (using Omron digital apparatus model 7200, Kyoto, Japan) and obtained random blood sugar (RBS) by glucometer (SD diagnostics, Korea).

#### Diagnosis confirmation camp

Within one week of a set of screenings performed by ASHAs through home-visits, a diagnosis confirmation camp was conducted by the study supervisor to obtain a second set of blood pressure readings and a random blood glucose assessment. The supervisor would advise on three key life-style interventions, as appropriate a) smoking cessation and avoidance of other tobacco use, b) consuming fewer salt containing foods c) Consuming more of fruits, vegetables and less of fried foods. This camp was operated during screening time-period in each slum. All at-risk individuals would be advised further follow ups at designated public health facilities. Home visits were conducted by the supervisor in case of non-attendance in the camp. Supervisors also visited households to collect information about asset ownership.

#### Identification of “high-risk” individuals

All “high risk” individuals who are identified during the home-based screening by ASHA and diagnosis confirmation camp by supervisors were referred to a physician for risk stratification decision made based on standard operational definitions of elevated blood pressure and blood sugar as defined above. Our data collection tool also has an inbuilt decision support system for stratification decision. To ensure quality-check, supervisors used to cross-check 10% of blood pressure measurements, tobacco and previously known NCD questions. All such high-risk individuals were advised about tobacco cessation, dietary modification, and increase in physical activity as appropriate. They were provided a referral-slip with previous blood pressure and blood sugar values to facilitate decision making at the nearest UPHC.

#### Linkage for treatment initiation and continuation at UPHC

The “high risk” individuals identified and referred by study supervisors were free to seek care from either the nearest UPHC or any other source of their choice. In UPHC, they were evaluated by a physician in the designated non-communicable disease clinic. The physicians were trained to follow simple therapeutic algorithms for treatment initiation, optimization and continuation for hypertension and diabetes. The physicians would preferably choose from available drugs at UPHC (Losartan (Angiotensin receptor blocker), Amlodepine (calcium channel blocker), Hydrocholorthiazide (diuretic), Metformin (Biguanide), Glimiperide (sulphonylurea), low-dose Aspirin, and Atorvastatin). All treatment decisions (Initiation, escalation, de-escalation of drug therapy) were recorded by the study physician in a NCD register available at UPHC for the study. The data of the NCD register was updated weekly into a designated data collection software by study supervisors. Periodic data quality checks were done by study investigators.

#### Follow-up visits by ASHAs and outcome classification

Subsequent to initial screening, ASHAs did home-visits, once in every two months to reinforce linkages to public health facilities and adherence to drug-treatment. Each visit would last for 30 to 45 minutes, and during each visit ASHA would perform a) review of baseline risk assessment; b)adherence to prescribed interventions; c) discussion about CVD prevention; d) measurements of blood pressure and blood sugar levels. Activities in these visits included counseling for at-risk individuals about risk factor modification, checking their preventative medical therapy, and advising them to make an appointment to visit a health facility, if required.

## Study Size

Main objective of the study is to explore feasibility of CHW based health promotion for reduction in CVD risk factors among those at high risk. Prevalence of such individuals at high risk is around 20 to 25%. There are around 350 notified slums in Bhopal consisting of 200 to 400 households per slum. Average household size is around 5 and thus there would be around 1000 to 2000 persons of which around 50-60% (500 to 1200) would be above 30 years. We have selected 16 slums randomly from within 10 km radius from our Institute. From these clusters ASHAs (CHWs) were asked to enroll all adults from households of catchment area or from 250 households whichever is lesser. This was done in order to screen around 7000 persons of above 30 years of age of which 1400 to 1750 were expected to be at high risk. This size of at high risk individuals would provide sufficient base to test feasibility of health promotion measures as well as its magnitude.

### Statistical Analysis

We have developed a data-collection tool based on CommcareHQ platform. Entire data collection activity was paperless. We have used tablet phone-based survey software for data collection. This facilitated real time, accurate data entry and saved on time and also it is environment friendly.

Commcare is a platform which was used for designing registration and follow up modules and forms. CHWs were given android mobile phones where they installed Commcare application. Initial registration was done by CHW after due consent. Then follow up information like measurement and BP readings were subsequently entered in follow up forms. This platform provides facility for tracking of participants through case management tool. CHWs were provided case list where they can search the enrolled participant by UID/name. Once patients were confirmed then this tool was used to assign color coded follow up priority. This color coding was based on BP and glycaemic control status of patients. It also had supervisor module in which supervisor was supposed to cross check and validate data of selected participants. For monitoring of project activities, XLS dashboard was also configured.

Data from the Commcare interface was exported as excel spreadsheet and checked for inconsistencies/missing information etc and cleaned accordingly. Information about age, years of education, BMI, WC, SBP, DBP, RBS will be recorded as continuous variables. Gender, marital status, current or ever tobacco use, alcohol use, prior HTN or DM, past manifest CVD were recorded as dichotomous or ordinal variables. These variables are a part of baseline survey. Time-logs from Commcare database were used to determine operational indicators such as screening-confirmation and confirmation-physician visit interval. Wealth Index was constructed by using principal component analysis of assets owned by each household. All data analysis was done using SPSS software (IBM SPSS Statistics for Macintosh, Version 26.0. Armonk, NY: IBM Corp.). Descriptive statistics are presented as means and standard deviations (or median and interquartile ranges if the data are not normally distributed) for continuous variables, and counts and percentages for categorical variables. As the individuals will be clustered within the household, we will be using multilevel modelling to evaluate the relationship between individual level exposures and outcomes of interest. We will also compute and compare the variable values at baseline and end line using paired t test or Wilcoxan signed rank test as appropriate.

Table-2 summarizes objective wise outcomes of the study and Table-3 describes logical framework of the study. Figure-1 shows flow diagram of the study procedures and Figure-2 shows geographic locations of the selected cluster along with geographic boundary.

**Table 1:** Interventions aimed at primary prevention of cardiovascular diseases in low and middle-income countries.

| First Author | Year of publication | Country / Region | Study type | Follow-up duration | Participants | Interventions | Outcome |
| --- | --- | --- | --- | --- | --- | --- | --- |
| *Randomized control trials involving community health workers* |  |  |  |  |  |  |  |
| Jafar T (30) | 2020 | Bangladesh, Pakistan, Sri Lanka | Cluster randomized controlled trial | 24 months | 2645 adults (1330 in intervention group and 1315 in control group) | Trained CHWs performing home visits for counselling on CVD risk factors and measuring BP | Mean SBP reduction was 5.2 mm Hg greater in intervention group (95% CI, 3.2 to 7.1, $P < 0.001$). All-cause mortality in intervention group was 2.9% and in control group was 4.3% |
| Peiris D (31) | 2019 | India | Stepped wedge, cluster randomized controlled trial | 24 months | Individuals in control arm were 4294 and in intervention arm were 4348 | CVD risk assessments by CHWs at household level in villages. Electronic referral and decision support (PHCs doctors). Follow up system. | No significant difference in outcome apart from minor increase in self-reported physical activity |
| Joshi R (32) | 2019 | India | Cluster randomized trial | 12 months | 28 villages in 3 states (2312 households with 3261 participants) | Trained CHWs giving education on CVD risk reduction and performing monitoring of risk factors | No change in SBP but adherence to anti-hypertensive drugs better in intervention arm in comparison to control arm (74.9% vs 61.4%, $P=0.001$) |
| Neupane D (33) | 2018 | Nepal | Open-label, cluster randomized trial | 12 months | 1638 (939 in intervention arm and 699 in control arm) | Female Community Health Volunteers (FCHV) performed home visits for lifestyle counselling and monitoring BP | Reduction in mean SBP in intervention arm in comparison to control arm in all groups (normotensive, pre-hypertensive and hypertensive) |

| *Community based primary care programs involving community health workers* |  |  |  |  |  |  |  |
| --- | --- | --- | --- | --- | --- | --- | --- |
| Gaziano T (34) | 2015 | Bangladesh, Guatemala, Mexico, South Africa | Observational study | 1.5 months | 4046 adults from community (Bangladesh=843 Guatemala=956 Mexico=1030 South Africa=1217)<br>42 CHWs | CHWs performing screening and predicting cardiovascular disease risk compared to physicians/nurses | Agreement between CHWs and physicians risk scores was 96.8% (weighted K=0.948, 95% CI 0.936-0.961). CHWs predicted 6% of participants had 5 year CVD risk greater than 20% |
| Reininger B (35) | 2015 | Mexico (US-Mexico border) | Observational study | 4 years | 1438 participants | Counselling by CHWs and Radio/TV messages on diet and physical activity | High association between meeting physical activity guidelines and exposure to CHWs and Radio/TV messages (adjusted OR=3.83,P=0.009) |
| *Facility based programs / interventions involving care-coordinators or health educators* |  |  |  |  |  |  |  |
| Kavita G (36) | 2020 | India | Quasi experimental study | 12 months | Nurses=16<br>Primary prevention in HTN Patients=402 (40 years and above)<br>Secondary prevention in CABG/PTCA patients=500 (Intervention=250 Control=250) | Counselling and CVD risk assessment by nurses in OPD of tertiary healthcare centre of Northern India | High agreement between nurses and investigator (k=0.84)<br>In primary prevention group, 70% in lower risk category as compared to 60.6% in baseline<br>In secondary prevention group, medication adherence score in intervention group was 7.6 compared to 5.9 in control group (P<0.01) |
| Shrivastava U (37) | 2017 | India | Randomized Controlled Trial | 6 months | 310 employees of worksites from Delhi and National Capital region | Sessions and training on healthy living, diet and physical activity by health professionals | Decrease in weight, subcutaneous fat and metabolic risk factors in intervention group |
| Jamal SN (38) | 2016 | Malaysia | Randomized Controlled Trial | 9 months | 194 obese and overweight university employees (intervention arm=97 control arm=97) | Group Support Lifestyle Modification (GSLiM) through self-monitoring, cognitive behaviour session, exercise and dietary change counselling. | GSLiM programme resulted into improvement in weight loss, weight self-efficacy, social support and quality of life in intervention arm vs control arm |
| Ray M (39) | 2016 | India | School based survey | 2 months | 2995 students (14-16 years) from 20 schools | School based CVD risk factors educational program through presentations by health educators | Improvement in post test score on CVD prevention and associated risk factors |
| **Ongoing studies** |  |  |  |  |  |  |  |
| Lijing Y (NCT01259700) | Ongoing | China | Community based cluster randomized trial | 2 years | 10,000 individuals | CVD risk reduction and salt reduction educational interventions | Primary outcome is blood pressure reduction |
| Prabhakaran D (NCT01212328) | Ongoing | India | Individual patient Randomized control trial | 42 months | 1120 patients with diabetes mellitus | Clinical care coordinator and decision support system | Multiple CVD risk control targets |
| Anchala R(28) (CTRI/2012/03/002476) | Ongoing | India | Facility based cluster-randomized trial | 12 months | 16 primary health center clusters (8 in each arm) | Physicians using a decision support system vs no such system | 4mm Hg reduction in systolic blood pressure |

NCT refers to NIH clinical trials registry number (clinicaltrials.gov); CTRI refers to Clinical trial registry of India number (www.ctri.in) All numbers indicate frequency (proportion) unless indicated otherwise.

**Table 2:** Objectives and Outcomes.

| **Primary Objectives** | **Key Outcome measure** |
| --- | --- |
| To assess prevalence of common modifiable risk factors (tobacco consumption, hypertension, diabetes, obesity, high salt consumption, physical inactivity etc) for CVDs in urban slums | Proportion of:<br>Current and former tobacco users<br>Current and newly diagnosed hypertensive<br>Current and newly diagnosed diabetics<br>Individuals overweight or obese<br>Individuals with abdominal obesity<br>Individuals with low physical activity scores<br>Individuals with high estimated salt consumption<br>Individuals with low intake of fruit & vegetables |
| To identify and train community health workers to screen individuals at a high risk for future cardiovascular events, and impart skills of self-care, risk reduction strategies and assessment of treatment adherence. | Number of CHWs trained<br>Change in CVD related health literacy and competence as determined by Pre-test and post test scores |
| To increase awareness about common modifiable risk factors using inter-personal communication. | Change in CVD related health literacy among the screened communities |
| **Secondary Objectives** | **Key Outcome measure** |
| To improve linkages of communities to public health delivery services | Proportion of individuals registered at NCD clinic and or Tobacco cessation clinic of public health delivery system. |
| To evaluate feasibility of community health worker-based adherence promotion strategies | Proportion of individuals accepting CHW visits<br>Proportion of completed CHW visits<br>Average time spent by CHW for each visit |

Those linked to Public health facilities are referent; OR=Odds ratio; ASHA=Accredited social health activist; HTN=Hypertension; DM=Diabetes; BMI=Body mass index; UPHC=Urban Primary health centre.

**Figure 1:**
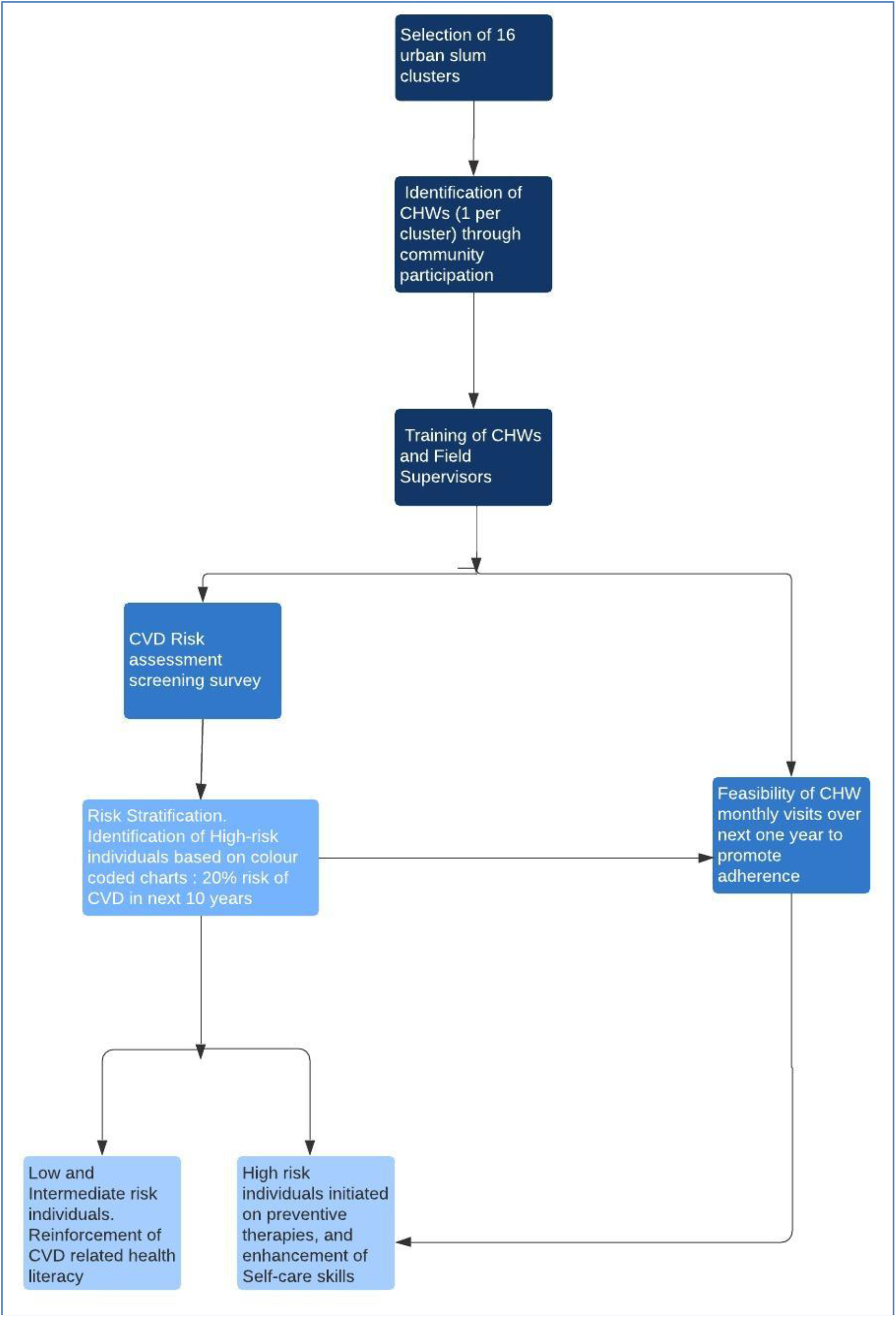
Flowchart showing study procedures.

**Figure 2:**
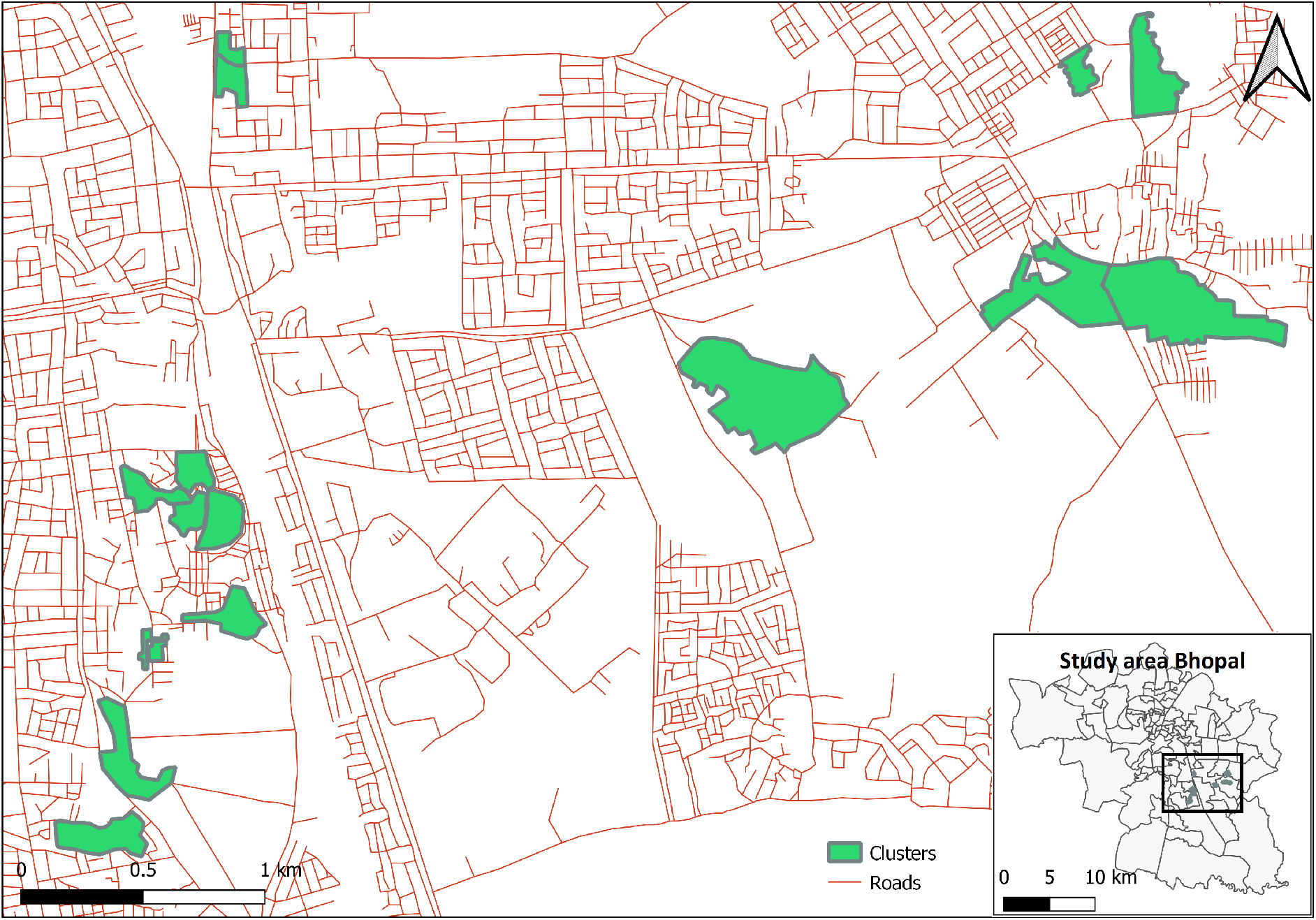
shows locations and area of selected clusters. Map of Bhopal city is shown in inset.

## Results of Baseline Survey

Between November 2017 and June 2018 a total of 6174 individuals of age 30 years or more, residing in 3424 households were screened. Of them 56.1% were women. This baseline survey was conducted by ASHA workers in a door to door fashion. They performed a socio-economic and behavioural assessment, performed anthropometry, and measured blood pressures and blood sugars. Of the 6174 individuals, visited at home, 4781 (77.43%) also attended a confirmation camp, conducted in the same clusters to confirm the high CVD risk status by a physician. (Table 4)

**Table 3:** The logical framework.

| **Objective** | **Rationale** | **Objectively verifiable indicators of achievement** | **Sources and means of verification** | **Assumptions** |
| --- | --- | --- | --- | --- |
| To assess prevalence of common modifiable risk factors (tobacco consumption, hypertension, diabetes, obesity, high salt consumption, physical inactivity etc) for CVDs in urban slums | Estimate of background prevalence is necessary for future intervention | Frequency and proportion of risk factors as obtained in the screening survey | Screening records | Robust definitions, minimum interviewer and observer bias, calibrated instruments and minimal measurement error and interobserver variability. |
| To identify and train community health workers to screen individuals at a high risk for future cardiovascular events, and impart skills of self care, risk reduction strategies and assessment of treatment adherence. | CHW identified from within the communities of intervention need to be trained in CVD risk reduction skills. | Proportion of CHWs who belong to the community of intervention. Pretest and post test scores and incremental change in knowledge. | Residence proof documentation<br>Supervisor based skill observation (OSCE) | Sustainability of CHWs<br>Adequate uptake of training<br>Completion of training |
| To increase awareness about common modifiable risk factors using inter-personal communication. | Health education of the communities will facilitate change in perception of susceptibility as well as perceived benefits. | Pre and post intervention Knowledge, Attitude, Behavior and Practices Survey. | Verification of 10% random sample | 10% random sample is representative of total |

**Table 4:** Distribution of behavioural risk factors among study participants (n=6174)

| **Variable** | **Frequency (Percent)** |
| --- | --- |
| No. of households surveyed | 3424 |
| No. of individuals registered | 6174 |
| Gender |  |
| Men | 2707 (43.8) |
| Women | 3466 (56.1) |
| Transgender | 1 (0.1) |
| Age [Mean (SD)] | 44.55 (12.27) |
| 30 - 44 years | 3355 (54.3) |
| 45 – 60 years | 1864 (30.2) |
| 61 – 74 years | 795 (12.9) |
| 75 years or more | 160 (2.6) |
| Education |  |
| No formal schooling | 1971(31.9) |
| 1 to 4 years | 446 (7.2) |
| 5 to 10 years | 2775 (44.9) |
| 11 years or more | 982 (15.9) |
| Marital status |  |
| Currently married | 5470 (88.6) |
| Not currently married | 704 (11.4) |
| No. of participants who attended confirmation camp | 4781 (77.43) |

Key behavioural risk factors in the population were smoking or tobacco use (Current users 38.8%), and lack of physical activity. Tobacco use was much more common as compared to smoking. Of all current users of these products, 11.7% smoked, 79.7% chew tobacco and only 8.5% used both the products. Mean duration of usage of such products was 21.9 years. Given the mean age of entire study population of 44. 5 years, individuals were consuming tobacco for about half their lives. (Table 5)

**Table 5:** Distribution of behavioural risk factors among study participants (n=6174)

| Variable | All | Women | Men | p-value |
| --- | --- | --- | --- | --- |
| **Total number** | **6174** | **3466** | **2707** |  |
| **Smoking or tobacco use** |  |  |  |  |
| Never used | 3640 (59.0) | 2682 (77.4) | 975 (35.4) |  |
| Past user | 141 (2.3) | 41 (1.2) | 100 (3.7) |  |
| Current user | 2393 (38.8) | 743 (21.4) | 1650 (61.0) | <0.0001 |
| **Among current and past users** |  |  |  |  |
| Smoking alone | 296 (11.7) | 6 (0.8) | 290 (16.6) |  |
| Oral alone | 2038 (79.7) | 773 (98.6) | 1254 (71.3) |  |
| Both | 218 (8.50) | 5 (0.6) | 213 (12.2) | <0.0001 |
| **Smoking or tobacco use duration** |  |  |  |  |
| Mean (years)* | 21.9 (14.37) | 21.1 (16.2) | 22.2 (13.5) | 0.08 |
| Less than 10 years | 643 (25.4) | 280 (35.7) | 363 (20.7) |  |
| 11 to 20 years | 735 (29.0) | 171 (21.8) | 564 (32.2) |  |
| 21 years or more | 1156 (45.6) | 333 (42.5) | 823 (47.0) | <0.0001 |
| **Alcohol use** |  |  |  |  |
| Never used | 4959 (80.3) | 3314 (95.6) | 1644 (60.7) |  |
| Ever used | 1215 (19.7) | 152 (4.4) | 1063 (39.3) | <0.0001 |
| **Alcohol use frequency among ever users** |  |  |  |  |
| Less than once a month | 342 (28.1) | 80 (52.6) | 262 (24.6) |  |
| Few days in a month | 434 (35.7) | 45 (29.6) | 389 (36.6) |  |
| 1-4 days in a week | 201 (16.5) | 13 (8.6) | 188 (17.7) |  |
| 5-6 days a week | 28 (2.3) | 1 (0.7) | 27 (2.5) |  |
| Daily | 210 (17.3) | 13 (8.6) | 197 (18.5) | <0.0001 |
| **Physical Activity / Lifestyle** |  |  |  |  |
| Non-sedentary | 1476 (23.9) | 623 (18.0) | 853 (31.5) |  |
| Sedentary | 4697 (76.1) | 2843 (82.0) | 1854 (68.5) | <0.001 |

All values above indicate number (percentage of total) except where indicated
\* Indicated mean (SD)

Sedentariness was another behavioural risk factor, seen in 76.1% of the study population. This was measured as a leisure time sedentariness, which was higher in women as compared to men. A total of 12.3% of the study population had known hypertension, and of these only 46.7% were controlled. Another 15% of all individuals had a newly detected hypertension. While prevalence of pre-existing hypertension was higher in women, prevalence of unknown hypertension was significantly higher in men. A total of 5.4% of the population had a pre-existing known diabetes, 57.5% of these being controlled. Another 3.1% of individuals had a newly detected diabetes mellitus. Prevalence of newly detected diabetes mellitus was also significantly higher in men. A total of 47.7% of all individuals had abdominal obesity, and 37.9% had a BMI higher than 25 kg/m2. Both abdominal obesity and high BMI state was more common in women as compared to men. (Table 6)

**Table 6:** Distribution of Biological Risk Factors (n=6174)

| **Variable** | **All** | **Women** | **Men** | **p-value** |
| --- | --- | --- | --- | --- |
| **Known disease history (n)** |  |  |  |  |
| Hypertension | 758 (12.3) | 514 (14.8) | 243 (9.0) | <0.001 |
| Uncontrolled (>140/90) | 404 (53.3) | 258 (50.2) | 145 (59.7) | 0.015 |
| Controlled (<140/90) | 354 (46.7) | 256 (49.8) | 98 (40.3) |  |
| Diabetes | 333 (5.4) | 196 (5.7) | 137 (5.1) | 0.305 |
| Blood sugars measured | 294 | 179 | 115 |  |
| Uncontrolled (RBS >200mg/dL) | 125 (42.5) | 87 (48.6) | 38 (33.0) | 0.031 |
| Controlled (RBS <200mg/dL) | 169 (57.5) | 92 (51.4) | 77 (67.0) |  |
| Ischemic Heart Disease | 85 (1.4) | 36 (1.0) | 49 (1.8) | 0.010 |
| Cerebrovascular disease | 65 (1.1) | 24 (0.7) | 41 (1.5) | 0.002 |
| **Previously not known hypertension** | **5416** | **2952** | **2464** |  |
| Normal (BP <120/80) | 2555 (47.2) | 1634 (55.4) | 921 (37.4) |  |
| Pre-hypertension (<140/90) | 2048 (37.8) | 949 (32.1) | 1099 (44.6) |  |
| Newly detected hypertension | 813 (15.0) | 369 (12.5) | 444 (18.0) | <0.001 |
| **Previously not known Diabetes** | **5841** | **3270** | **2570** |  |
| **Blood sugars in not known Diabetes** | **4486** | **2698** | **1788** |  |
| Normal Random sugar (<140mg/dl) | 3744 (83.5) | 2296 (85.1) | 1448 (81.0) |  |
| Impaired glucose (140-199mg/dL) | 591 (13.2) | 326 (12.1) | 265 (14.8) |  |
| Newly detected Diabetes (>200mg/dl) | 151 (3.4) | 76 (2.8) | 75 (4.2) | 0.00075 |
| **Average blood pressure values among individuals with hypertension\*** |  |  |  |  |
| *Previously known hypertension (n=757; 514 women, 243 men)* |  |  |  |  |
| Systolic blood pressure | 142.4 (23.1) | 142.0 (23.8) | 143.1 (21.7) | 0.541 |
| Diastolic blood pressure | 84.4 (13.7) | 83.3 (13.4) | 86.9 (13.9) | <0.001 |
| *Newly detected hypertension (n=813; 369 women, 444 men)* |  |  |  |  |
| Systolic blood pressure | 152.1 (15.9) | 153.7 (15.5) | 150.8 (16.1) | <0.001 |
| Diastolic blood pressure | 92.5 (10.7) | 90.2 (11.1) | 94.5 (9.9) | <0.001 |
| *Previously known Diabetes (n=315; 194 women, 121 men)* |  |  |  |  |
| Random Blood Sugar | 200.0 (84.0) | 208.0 (83.4) | 187.6 (83.8) | 0.883 |
| *Newly detected Diabetes (n=151; 76 women, 75 men)* |  |  |  |  |
| Random Blood Sugar | 280.3 (64.7) | 278.9 (61.3) | 283.4 (72.8) | 0.042 |
| **Waist circumference(n)** | **6162** | **3462** | **2700** |  |
| Mean Waist Circumference |  | 82.7(15.5) | 84.2(16.0) | <0.001 |
| Increased (>80cm in women or >90cm in men) | 2940 (47.7) | 2068 (59.7) | 872 (32.3) | <0.001 |
| **Body mass index (n)** | **4780** | **2877** | **1903** |  |
| Mean Body Mass index | 24.0(4.8) | 24.7(5.1) | 23.0 (4.2) | <0.001 |
| Less than 18.5 | 551 (11.5) | 306 (10.6) | 245 (12.9) |  |
| 18.5 to 24.9 | 2419 (50.6) | 1296 (45.0) | 1123 (59.0) |  |
| 25 to 29.9 | 1328 (27.8) | 896 (31.1) | 432 (22.7) |  |
| More than 30 | 482 (10.1) | 379 (13.3) | 103 (5.4) | <0.001 |

\* All average blood pressure values are in mean (SD). Remaining values are numbers (percentage of total)

The gender-based differences in risk factors were significantly different and these are represented in Figure 3.

**Figure 3:**
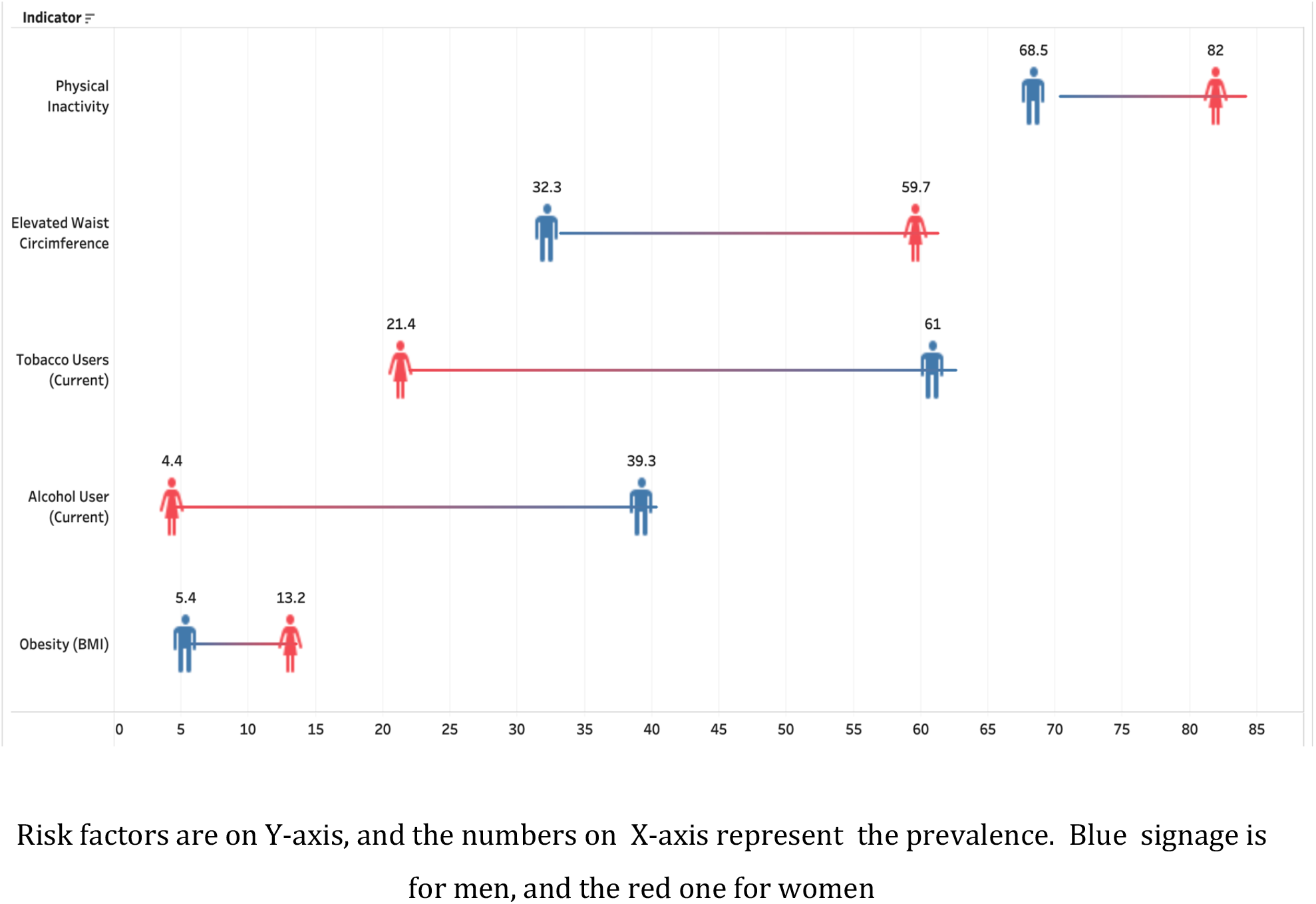
Prevalence of Behavioural and Biological risk factors by Gender (n=6174)

Further analysis as described in statistical analysis section is in process and will be shared in separate manuscript.

## Discussion

While many CHW based interventions have been shown to be beneficial in promoting childhood immunization uptake, initiation of breast-feeding and improving pulmonary TB cure rates (22-24), there is a paucity of literature on their role in cardiovascular disease risk reduction, especially in low-and-middle income countries (LMICs). As per a systematic review and meta-analysis conducted by Widmer et al. on the 51 studies involving digital health interventions (smart phones, mobile based applications, telemedicine, etc.) from 1990 to 2014, there was substantial decline in CVD outcomes after using such interventions (RR=0.61,P=0.002,I^2^=22%) (25). Pennant and co-workers performed a meta-analysis of cardiovascular disease risk reduction programs from 1970 to 2008. In this meta-analysis, all the 36 identified programs were from high-income settings (26).Overall there was a 0.65% absolute reduction in 10-year CVD risk, with many interventions achieving as much as a 1% 10-year risk-reduction. None of the programs, however, achieved a significant reduction in mortality. As per the systematic review and meta-analysis carried by Palmas et al. in 2015, CHW based interventions reduce the A1c levels in the community (27). Another systematic review of CHW based intervention trials (28),identified 10 trials which targeted hypertension, diabetes mellitus, or CVDs. Nine of these were from high income countries. These trials demonstrated that CHW interventions can increase knowledge levels, improve follow up rates, and achieve better glycemic or blood pressure control. A systematic review of trials in LMICs (2017) depicted that CHWs based implementation of CVD reduction has promising potential specially with respect to domains like control of hypertension, diabetes and tobacco consumption (29).

We reviewed published and ongoing studies aimed at cardiovascular disease risk reduction from LMICs. A recent cluster randomized controlled trial (Feb,2020) performed in rural districts of Bangladesh, Pakistan and Sri Lanka depicted that by involving trained CHWs for counselling adults on CVD risk factors and monitoring BP, a significant reduction in mean SBP and all-cause mortality in intervention group is possible (30). A stepped wedge cluster randomized controlled trial conducted in India (2019) showed null difference in intervention and control arm with respect to CVD risk assessment by the CHWs at the level of village households with incorporation of clinical decision support system by PHC doctors for reduction in CVDs (31). Another cluster randomized trial in India (2019) involving CHWs for delivering CVD risk reduction education accompanied by monitoring of risk factors, depicted no significant change in SBP but improved adherence to anti-hypertensive drugs in intervention arm compared to control arm (74.9% vs 61.4%, P=0.001) (32). As per an open label cluster randomized trial carried in Nepal (2018), a significant reduction in mean SBP (−4.9 mm Hg, 95% CI at -7.78 to -2, P=0.001) was evident in intervention group which included Female Community Health Volunteers (FCHVs) delivering lifestyle counselling and monitoring BP at the level of community (33).

As per a study conducted by Gaziano et al., CHWs were found to be proficient not only in screening adults for CVDs but also on predicting the CVD risk scores like health professionals (weighted K=0.948, 95% CI 0.936-0.961) (34). Another study highlighted the role of CHWs counselling and media (radio/television) messages in improving the physical activity levels in community members of Mexico through the Community Wide Campaign (CWC) named as *Tu Salud, Si Cuenta* (35). A few other educational interventions at health-care facilities (36), workplaces (37,38) and schools (39) which do not involve CHWs in the true sense, have also reported an improvement in risk factor levels and knowledge. (Table 1)

There are evidence-practice gaps in CVD prevention, which operate at various levels.(40) Individuals who are screened for hypertension and diabetes mellitus in the community are largely asymptomatic. They need to be aware of future risks of elevated blood-pressure and blood glucose, convinced that these risks need to be reduced, and take supervised affirmative measures for change in risk behaviours and uptake of life-long preventive therapy. Such life-long preventive therapy has a complex behavioural construct as it requires certain activities to be reduced (such as tobacco and alcohol use, consumption of excess salt, and calories) and others to be enhanced (such as physical activity, fruits/vegetables and adherence to drug therapy). While availability and affordability of CVD preventive therapies, especially in Low- and Middle-income Countries (LMICs) are believed to be a key impediment in optimal risk factor control, these may not be the only factors. Despite a much better availability and affordability of anti-hypertensive medication in high income countries, proportion of individuals with optimal control is only marginally better (36% in high income vs 23% in low-income countries).(41) This implies that additional patient and health-care provider level factors (knowledge, attitude, beliefs and practices), and health-system barriers (infrastructure, access, quality) are equally important.(42)

Under NPCDCS, population-based screening (PBS) has been recently introduced in India. The current study was designed in context of a community health worker (CHW) led screening and preventive therapy facilitation initiative for CVD prevention in urban slum communities. Linkage to public health primary care facilities was a key outcome indicator for this initiative. In the current study we have tried to answer two questions; first, who among the high risk individuals do not get linked (predictors of non-linkage), and second, why they do not get linked (barriers to non-linkage). Thus the prediction research question was to determine that among high CVD risk individuals, presence of which demographic, socio-economic, CVD risk-factor and environmental characteristics (as compared to their absence) are associated with non-linkage to public sector primary care facilities. The barrier research question was that among high CVD risk individuals, which individual, health-system, and societal determinants were more likely to be associated with non-linkage behaviour.

We have also presented baseline summary of socio-demographic characteristics individuals screened, presence of behavioral and biological risk factors for CVD and status of blood pressure and glycemic control

## Strengths and Limitations

Strength of this study lies in the settings in which it is conducted. The study was implemented through CHWs affiliated to catchment area of Urban Primary Health Centres. A national health program (NPCDCS) envisages population-based screening through these workers, and further linkage to public health facilities. Our study thus provides insights into feasibility of CHW based screening, detection, and treatment initiation interval and also barriers which leads to non-initiation or discontinuation after treatment. Another strength lies in data collection and decision support system tools used in study. A mobile phone-based data collection system deployed on CommCare enabled real-time data availability which was necessary for prioritizing CHW based follow up visits. We have implemented this study through CHWs who were part of existing health care delivery system and thus had routine competing priorities for tasks to be done in this project. Thus, project tasks may have been on low priority. This can be considered as strength (tells us about real field picture) as well as limitation (uncertainty about actual performance when such kind of program is scaled up as mainstream program). Another limitation lies in the fact that, out of 6178 individuals initially registered by ASHA, approximately 1800 did not turn up to confirmation camps and their second blood pressure reading or blood sugar reading was not available for confirming ‘high risk’ status. This partial response may have affected our study results. However, this number was reduced to minimum by considering their only blood pressure reading at home for classification and including all self-reported persons with hypertension and diabetes as ‘high risk’.

## Conclusions

We need robust mechanisms to monitor adherence especially when a large number of individuals are likely to be screened and treated in public sector.(43) We also need an efficient health-system that ensures continual access to medication, with minimum disruption of occupational priorities. Recent guidelines for hypertension, and higher CVD risk in South-Asians, advocates a more aggressive management of hypertension and diabetes mellitus.(44) Various systematic reviews have recorded numerous successful interventions, to overcome barriers at individual, family, community, provider, and health-system levels.(45) Interventions that addressed barriers at multiple-levels were more successful than the interventions that focused on a single or fewer barriers.(46) Some of the cost-effective solutions could include improved information and behavior change measures by community health workers, reinforcements by family and providers, improved drug packaging, accessibility, and monitoring mechanisms at the health-system levels.

## Data Availability

Dataset used in this manuscript can be made available to interested researchers upon request to corresponding author. It is not being deposited in public repository since final analysis is in process and this manuscript mainly details study protocol. Dataset will be deposited in public repository with manuscript sharing final results.

## Author contributions-

Conceptualization: [Rajnish Joshi, Abhijit Pakhare, Sanjeev Kumar], Methodology: [Rajnish Joshi, Abhijit Pakhare, Sanjeev Kumar]; Formal analysis [Rajnish Joshi, Abhijit Pakhare, Ankur Joshi]; visualization: [Yogesh Sabde, Abhijit Pakhare, Anuja Lahiri]; investigation: [Sagar Khadanga, Vaibhav Ingle, Shubham Atal, Akash Ranjan, Neelesh Shrivastava, Anuja Lahiri]; Supervision: [Rajnish Joshi]; Writing - original draft preparation: [Rajnish Joshi, Anuja Lahiri]; Writing - review and editing: [All authors]; Funding acquisition: [Rajnish Joshi]

## Ethics approval-

The study design was approved by the Institutional Human Ethics Committee of All India Institute of Medical Sciences, Bhopal (Ref: IHEC-LOP/2017/EF00045)

## Informed consent-

Participant Information Sheet in Hindi language was provided to each participant. All participants provided written informed consent prior to initiation of any study procedures.

## Data Availability Statement

Dataset used in this manuscript can be made available to interested researchers upon request to corresponding author. It is not being deposited in public repository since final analysis is in process and this manuscript mainly details study protocol. Dataset will be deposited in public repository with manuscript sharing final results. Acknowledgements: We are thankful to Chief Medical and Health Officer, Bhopal, Medical Officers of Urban Primary Health Centres at Barkheda Pathani and Saibaba Nagar, Bhopal and Program Officers concerned with ASHAs (CHWs) who have facilitated implementation of this study. Also, we are thankful to all ASHAs who have executed the study.

